# Reduction in the risk of mpox infection after MVA-BN vaccination in individuals on HIV pre-exposure prophylaxis: a Spanish cohort study

**DOI:** 10.1101/2023.05.30.23290712

**Authors:** Mario Fontán-Vela, Victoria Hernando, Carmen Olmedo, Ermengol Coma, Montse Martínez, David Moreno-Perez, Nicola Lorusso, María Vázquez Torres, José Francisco Barbas del Buey, Javier Roig-Sena, Eliseo Pastor, Antònia Galmés Truyols, Francisca Artigues Serra, Rosa María Sancho Martínez, Pello Latasa Zamalloa, Olaia Pérez Martínez, Ana Vázquez Estepa, Amós José García Rojas, Ana Isabel Barreno Estévez, Alonso Sánchez-Migallón Naranjo, Jaime Jesús Pérez Martín, Pilar Peces Jiménez, Raquel Morales Romero, Jesús Castilla, Manuel García Cenoz, Marta Huerta Huerta, An Lieve Dirk Boone, María José Macías Ortiz, Virginia Álvarez Río, María Jesús Rodríguez Recio, María Merino Díaz, Belén Berradre Sáenz, María Teresa Villegas-Moreno, Aurora Limia, Asuncion Diaz, Susana Monge, Spanish MPOX vaccine effectiveness study group

## Abstract

**Objectives:** To assess the effectiveness of at least one-dose of the Modified Vaccinia Ankara-Bavaria Nordic (MVA-BN) smallpox vaccine, administered pre-exposure, against mpox virus (MPXV) infection in persons receiving pre-exposure prophylaxis for HIV (HIV-PrEP).

**Design:** Retrospective cohort, constructed by deterministic linkage of electronic health records.

**Setting:** 15 of 19 regions in Spain (>95% population), between July 12 and December 12, 2022

**Participants:** Men ≥18 years, receiving HIV-PrEP as of July 12 and with no previous MPXV infection or vaccination against mpox.

**Interventions:** On each day, we matched individuals receiving a first dose of MVA-BN vaccine pre-exposure and unvaccinated controls of the same age (±5 years) and region.

**Main outcome measures:** We estimated the risk of laboratory-confirmed infection with MPXV using a Kaplan-Meier estimator and calculated risk ratios (RR) and vaccine effectiveness (VE=1-RR).

**Results:** We included 5,660 matched pairs, with a median follow-up of 62 days (interquartile range 24-97). Mpox cumulative incidence was 5.6 per 1,000 (25 cases) in unvaccinated and 3.5 per 1,000 (18 cases) in vaccinated; last case occurred at 63 and 17 days after enrolment, respectively. No effect was found during days 0-6 post-vaccination (VE -38.3; 95% confidence interval (95%CI): -332.7; 46.4), but VE was 65% in ≥7 days (95%CI 22.9; 88.0) and 79% in ≥14 days (95%CI 33.3; 100.0) after vaccination.

**Conclusion:** At least one dose of MVA-BN vaccine offered protection against mpox in a most-at-risk population. Because the incidence of mpox was decreasing shortly after the vaccination campaign began, we can only assess its effectiveness shortly after vaccination. Further studies need to assess the VE of a second dose and the duration of protection over time.

- **What is already known on this topic:**
  - MVA-BN vaccination is effective in producing an immune response against mpox virus (MPXV) and has shown clinical efficacy in animal models.
  - During the current mpox outbreak, observational studies have indicated that MVA-BN vaccines are effective in preventing MPXV infection in real-life.
  - No individual-based cohort study with proper implementation of causal inference methods, providing the highest quality of evidence with observational data, has been published to date.
- **What this study adds:**
  - The administration of one dose of MVA-BN vaccine in a high-risk population (men receiving HIV pre-exposure prophylaxis) reduced the risk of MPXV infection by 65% starting 7 days after the administration and by 79% starting 14 days after.
  - Effectiveness of MVA-BN vaccine was similar in population under 50 years, a group where childhood smallpox vaccination is rare in Spain.
  - This is the first study to estimate MVA-BN vaccine effectiveness in one of the countries most affected by the mpox outbreak in 2022, with a design and methods providing high quality evidence.

## Introduction

In early May 2022, an outbreak of mpox (formerly named monkeypox) emerged and rapidly spread through Europe, the Americas and other regions, with over 87,000 cases and 140 deaths notified by 111 countries one year later [1]. Spain is the country with the highest cumulative incidence in Europe and the third globally (only after the United States of America and Brazil), with over 7,500 notified cases [2]. In the current outbreak, person-to-person transmission has occurred predominantly through direct contact with skin lesions or with body fluids during sexual intercourse, or in other situations with continuous and prolonged close physical contact [3–6]. A high proportion of notified cases are men who have sex with men (MSM) [3,6–8].

Prevention and control measures for the current outbreak have included vaccination with Modified Vaccinia virus Ankara (MVA). MVA-BN (from Bavarian Nordic, BN, branded as JYNNEOS or IMVANEX in America and Europe, respectively) is a third-generation vaccine against smallpox containing a non-replicative live virus with a better safety profile [9]. At the beginning of the outbreak, MVA-BN vaccines were scarce and were prioritized as post-exposure prophylaxis in persons with known risk contact with an infected case, given within 4 days of the contact (or up to 14 days if there were no symptoms) [10]. With increasing availability, pre-exposure vaccination was recommended for persons with no previous vaccination against vaccinia virus at high risk of mpox, specifically, if they had high number of sexual partners (≥10 in the last year or ≥3 in the last 3 months), had been involved group-sex activities, or had a sexually transmitted infection (STI) diagnosed in the last month [11]. The recommended schedule is two doses administered ≥28 days apart, either subcutaneous (0.5 ml) or intradermal (0.1 ml). In Spain, the first dose of MVA as pre-exposure prophylaxis was started on 12 July 2022 and the second dose on 6 September 2022 [12].

Before the current outbreak, no estimates of clinical efficacy were available outside of animal models [13,14]. During the outbreak, estimates of vaccine effectiveness (VE), when administered pre-exposure, have been produced by the United Kingdom [15], the United States [16–19] and Israel [20], though only the latter was a cohort study, and had some methodological limitations [21]. The greatest difficulty for individual-based mpox VE studies has been the lack of a sampling frame of the population targeted for vaccination, needed to identify vaccinated and unvaccinated groups that are similar in risk practices and risk of mpox virus (MPXV) infection, independently of the probability of receiving a dose of MVA-BN vaccine.

Individuals receiving HIV pre-exposure prophylaxis (HIV-PrEP) are a well-identified population in Spain, due to HIV-PreP is only prescribed at hospital pharmacy services, are at high risk of MPXV infection and have been proactively targeted for pre-exposure MVA vaccination [22]. Our aim was to estimate the reduction in the risk of MPXV infection associated to the administration of at least one dose of MVA-BN vaccine pre-exposure in persons receiving HIV-PrEP in Spain.

## Methods

### Study design and setting

We constructed a retrospective cohort study by deterministic linkage of databases using any of three personal identifiers (national health system number, national identification number, and regional health system number). Spanish regions (Autonomous Regions) have the responsibility to deliver healthcare to the citizenship and manage vaccination programs. We collected data from 15 out of 19 regions, encompassing over 95% of the Spanish population: Andalusia, Asturias, Balearic Islands, Canary Islands, Castile and León, Castilla-La Mancha, Catalonia, Valencian Community, Extremadura, Galicia, Community of Madrid, Region of Murcia, Navarre, Basque Country, and La Rioja. The regions reported individual-level data from three data sources: (i) all diagnoses of MPXV infection during the outbreak; (ii) all MVA-BN vaccine-doses administered; and (iii) the list of individuals receiving HIV-PrEP as of 12 July 2022. Individual identifiers were pseudo-anonymized using a HASH algorithm, a deterministic unidirectional coding system that preserves anonymity while allowing linkage. The three data sources were linked centrally, to allow the curation of duplicates and identification of all vaccines and infections nationally, except for the Balearic Islands, Community of Madrid and Navarre, who sent the data cross-matched and completely anonymized.

### Specification of the target trial

Our observational study emulated a hypothetical target trial to estimate the effect of the administration of at least one dose of MVA-BN vaccine for the prevention of infection with MPXV in a most-at-risk population. The target trial would start on 12 July 2022, and the eligible population would be men older than 18 years, who were receiving HIV-PrEP on that date, and with no prior MPXV infection or vaccination since the beginning of the mpox outbreak in Spain.

In the target trial, eligible individuals would be randomly assigned to either the administration of a first dose of MVA-BN vaccine (regardless of the vaccine brand or the administration route) or to no administration of vaccine within strata defined by age and region. The outcome of interest would be the date of laboratory-confirmed MPXV infection.

### Emulation of the target trial

We emulated the target trial with the linked observational data, starting on 12 July 2022 and ending on 12 December 2022, when the first region extracted the data for the study. We excluded individuals with possible information errors, such as those that started HIV-PrEP before it was included in the National Health System (November 2019), those with missing date of infection and those with ≥3 doses of MVA-BN vaccine during the study period. Some regions did not have the information on sex within the HIV-PreP registry and it was assumed that all were males, since nearly all HIV-PrEP users (99.7%) are men [23]. To emulate the trial, on each day between 12 July and 12 December 2022, we identified individuals who met the eligibility criteria and classified them as either having or not having received a first dose of MVA-BN vaccine that day. Each vaccinated person was matched to a randomly selected control among eligible individuals who had not received any dose of vaccine up to that date. Exact matching was performed, with replacement, on age (± 5-years) and region. Vaccinated individuals could be matched as unvaccinated controls in the period up to one day before the first dose administration.

The outcome of the study was laboratory-confirmed MPXV infection, with the date of the event defined as the earliest between the date of symptoms onset or the date of laboratory-confirmation. For each matched pair, follow-up started on the day of administration of the first dose of MVA-BN vaccine and finished at the earliest of the date of event, death, or 12 December 2022. We followed a per-protocol approach to estimate VE, hence we censored both members of a matched pair when the control received the first dose of MVA-BN vaccine.

We performed a secondary analysis restricting to pairs in which both members were younger than 50 years, as a *proxy* of VE with no vaccination against smallpox during childhood. We were unable to assess the VE of two MVA-BN doses because no infection was registered after the administration of a second dose. Likewise, VE of only one dose of vaccine was equivalent to VE of at least one dose of vaccine (the main analysis).

### Statistical analysis

We computed the cumulative incidence (risk) curves of MPXV infection in vaccinated and unvaccinated groups using the Kaplan-Meier estimator [24]. We computed the Risk Ratio (RR) overall and at different points in time: for days 0-6 or ≥7 after the first dose administration or, alternatively, for days 0-13 or ≥14. To compute risk and risk ratios ≥7 and ≥14 days after vaccination, we used only matched pairs in which both individuals were still at risk at 7 (and 14) days after time zero. We computed 95% confidence intervals (95%CI) using non-parametric bootstrapping with 500 samples and applying the percentile bootstrap method [25]. We estimated vaccine effectiveness as VE = (1-RR)*100. Analyses were performed with R software version 4.1.2 (R Foundation for Statistical Computing).

To test the impact of the analytical approach [26], we conducted a sensitivity analysis to estimate the VE of one dose of MVA-BN vaccine using the full eligible population with time varying vaccination status, computing the number of events and time at risk by vaccination status, week, age group and region, and estimating adjusted Incidence Rate Ratio (IRR) with Poisson regression. The detailed methodology and results of this analysis are found in the Supplementary Material.

This study was approved by the Research Ethics Committee at the Institute of Health Carlos III (approval no. CEI PI 92_2022) and by the Research with Drugs Ethics Committee at the Community of Madrid (approval no. EV_MPOX-001).

## Results

### Description of study participants

We identified 10,449 eligible individuals, of which 5,920 (56,7%) received a first dose of MVA-BN vaccine and 2,014 (19.3%) two doses. Both the initial and the eligible population had similar characteristics (Supplement Table S1). We matched 5,660 (95.6%) individuals who received at least one dose of MVA-BN vaccine to the same number of controls who had not received vaccination against mpox up to that day (Figure 1). The unvaccinated group included 3,899 unique individuals, with a maximum number of repetitions of a matched control of 7. Censoring because the control received the first dose of MVA-BN vaccine occurred in 42.6% (n=2,412) of matched pairs.

**Figure 1.**
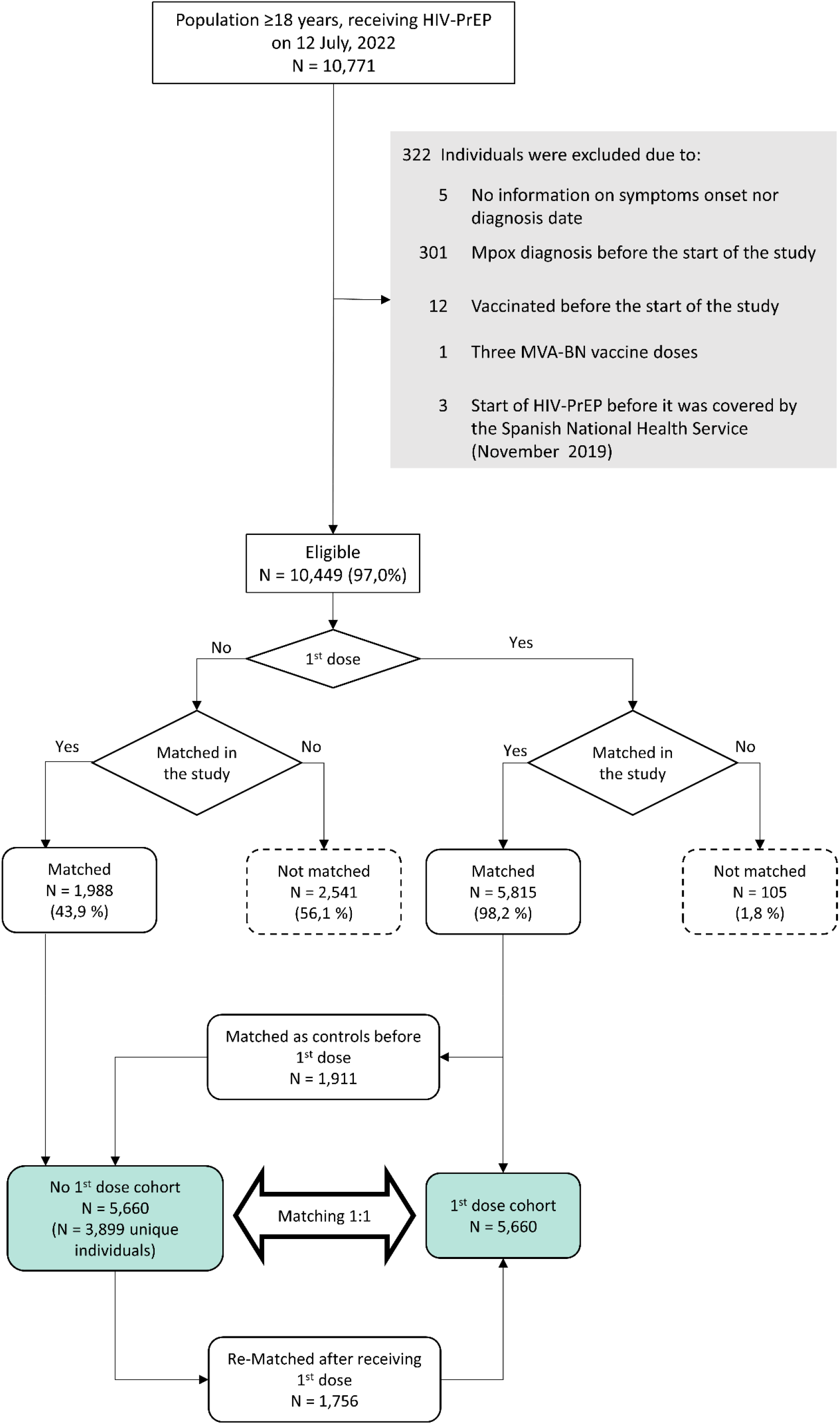
Sample selection flowchart. Abbreviation: HIV-PrEP, HIV pre-exposure prophylaxis.

Compared to the eligible population (Supplement Table S1), individuals in the matched sample had similar age (median 36 years, interquartile range [IQR]: 31-43). Virtually no individuals received a dose of smallpox vaccine during childhood in the matched sample (n=2 [0.0%]) compared to the eligible population (n=135 [1.3%]). No hospitalization, ICU admission or death was recorded in the matched sample, while 19 and 11 mpox cases were hospitalized in the initial and the eligible populations, respectively.

Table 1 shows the characteristics of the matched sample by vaccination status. The number of MPXV infections recorded in the matched sample was 43, with a higher number of cases among unvaccinated (25 vs 18 in the vaccinated).

**Table 1.** Characteristics of the individuals of the matched sample (N=11,320) by vaccination status

|  | Vaccinated<br>(n=5,660) |  | Unvaccinated<br>(n=5,660) |  |
| --- | --- | --- | --- | --- |
|  | n | % | n | % |
| **Age group, years** |  |  |  |  |
| 18-29 | 964 | 17.0 | 1,003 | 17.7 |
| 30-39 | 2,622 | 46.4 | 2,607 | 46.1 |
| 40-49 | 1,557 | 27.5 | 1,534 | 27.1 |
| ≥ 50 | 517 | 9.1 | 516 | 9.1 |
| **Childhood smallpox vaccination** |  |  |  |  |
| Yes | 1 | 0.0 | 1 | 0.0 |
| No | 44 | 0.8 | 48 | 0.8 |
| Unknown | 5,615 | 99.2 | 5,611 | 99.2 |
| **Autonomous Region** |  |  |  |  |
| Andalusia | 926 | 16.4 | 926 | 16.4 |
| Asturias | 37 | 0.7 | 37 | 0.7 |
| Balearic Islands | 361 | 6.4 | 361 | 6.4 |
| Canary Islands | 212 | 3.7 | 212 | 3.7 |
| Castile and León | 59 | 1.0 | 59 | 1.0 |
| Castilla-La Mancha | 59 | 1.0 | 59 | 1.0 |
| Catalonia | 2,121 | 37.5 | 2,121 | 37.5 |
| Valencian Community | 351 | 6.2 | 351 | 6.2 |
| Extremadura | 27 | 0.5 | 27 | 0.5 |
| Galicia | 252 | 4.5 | 252 | 4.5 |
| Community of Madrid | 830 | 14.7 | 830 | 14.7 |
| Region of Murcia | 129 | 2.3 | 129 | 2.3 |
| Navarre | 44 | 0.8 | 44 | 0.8 |
| Basque Country | 293 | 5.2 | 293 | 5.2 |
| La Rioja | 2 | 0.0 | 2 | 0.0 |
| MPXV infection |  |  |  |  |
| Yes | 18 | 0.3 | 25 | 0.4 |
| No | 5,642 | 99.7 | 5,635 | 99.6 |
| Mpox symptoms* |  |  |  |  |
| Yes | 18 | 100.0 | 25 | 100.0 |
| No | 0 | 0.0 | 0 | 0.0 |
| Hospitalization* |  |  |  |  |
| Yes | 0 | 0.0 | 0 | 0.0 |
| No | 18 | 100.0 | 25 | 100.0 |
| Admitted to ICU* |  |  |  |  |
| Yes | 0 | 0.0 | 0 | 0.0 |
| No | 18 | 100.0 | 25 | 100.0 |
| Death* |  |  |  |  |
| Yes | 0 | 0.0 | 0 | 0.0 |
| No | 18 | 100.0 | 25 | 100.0 |
| MVA-BN product |  |  |  |  |
| IMVANEX | 340 | 6.0 | - | - |
| JYNNEOS | 3,554 | 62.8 | - | - |
| Unknown | 1,766 | 31.2 | - | - |
| MVA-BN route of administration |  |  |  |  |
| Intradermal (0.1 ml) | 3,502 | 61.9 | - | - |
| Subcutaneous (0.5 ml) | 1,707 | 30.2 | - | - |
| Unknown | 451 | 7.9 | - | - |

\*Proportion is over the total number of MPXV infections

Figure 2a depicts the cumulative incidence in the vaccinated and unvaccinated groups during the study period. Median follow-up was 62 days (IQR: 24-97), with a maximum of 147 days. Cumulative incidence was 5.58 cases per 1,000 individuals in the unvaccinated compared to 3.46 per 1,000 in the vaccinated. Out of 25 mpox cases among unvaccinated, 8 (32%) and 13 (52%) cases were registered during the first 6 and 13 days respectively. Out of 18 mpox cases among vaccinated, 11 (61%) and 15 (83%) were reported in the same period. The last mpox case was registered after 63 days of follow-up in the unvaccinated and after 17 days in the vaccinated.

**Figure 2.**
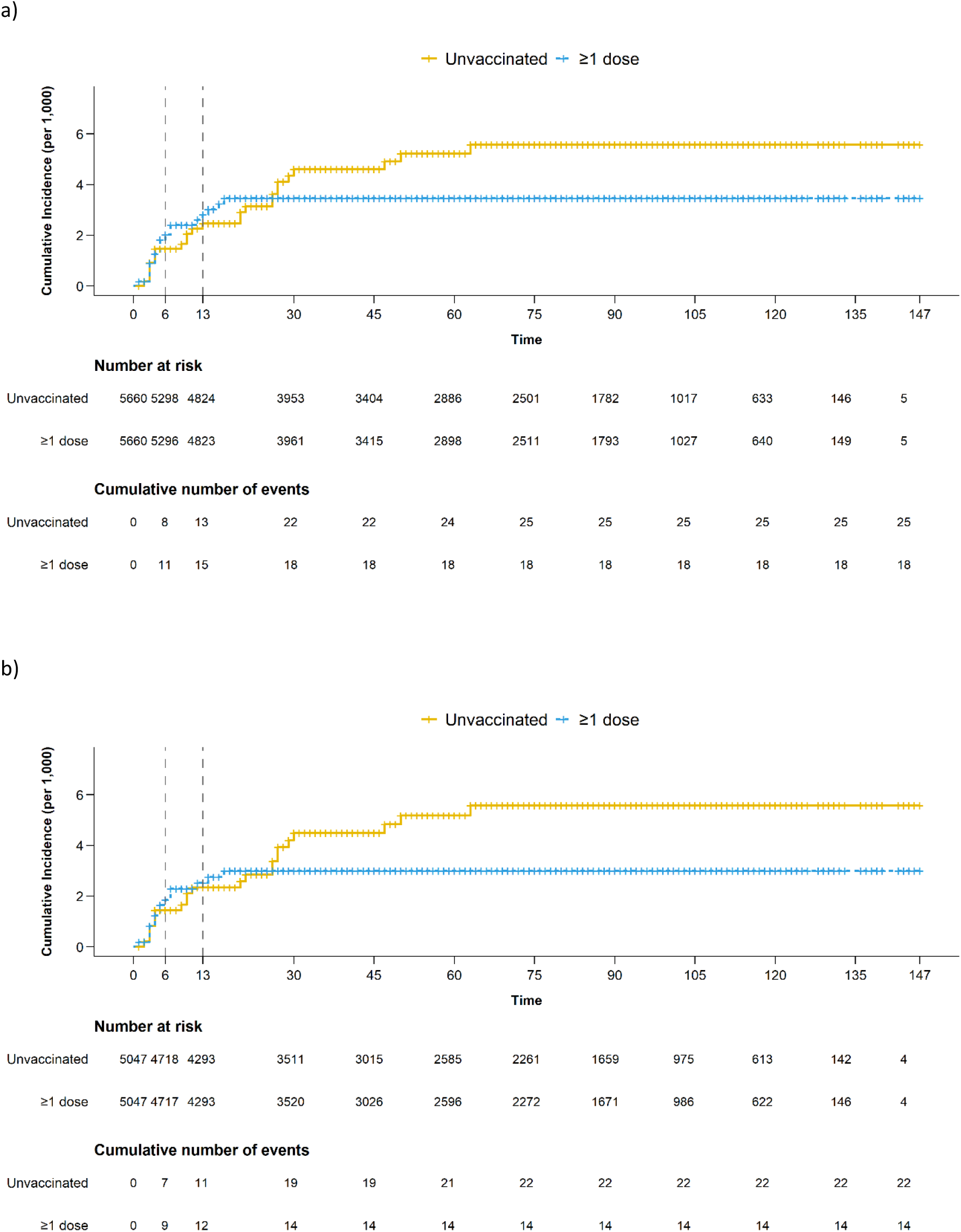
Estimated MPXV infection risk in the sample of individuals vaccinated with at least one dose of MBA-BN vaccines and matched unvaccinated controls, overall (a) and in individuals under 50 years of age (b)

### Effectiveness of one dose of MVA-BN

Since no event was registered after the administration of a second dose of MVA-BN vaccine, the VE of at least one dose of vaccine was equivalent to the VE of one dose.

During the study period, the overall estimated effectiveness of one dose of MVA-BN vaccine was 37.9% (95%CI -24.4; 69.1). During the first 6 and 13 days, the estimated VE was -38.3% (95%CI -332.7; 46.4) and -14.1% (95%CI -199.7; 47.9), respectively, showing a non-statistically-significant higher risk of MPXV infection in the vaccinated group in the immediate days after vaccine administration. At ≥7 days post-vaccination, the estimated VE was 65.0% (95%CI 22.9; 88.0), and it increased up to 79.3% (95%CI 33.3; 100.0) at ≥14 days.

### Results from secondary and sensitivity analyses

We restricted the analysis to 5,047 matched pairs in which both individuals were under 50 years of age. Among vaccinated individuals, 14 mpox cases were registered compared to 22 cases among the unvaccinated. The last case of mpox was registered 63 days after the enrollment in the unvaccinated and after 17 days in the vaccinated. The risk of MPXV infection was higher in unvaccinated individuals (5.6 per 1,000) than in the vaccinated (3.0 per 1,000) (Figure 2b and Table 3). VE from 7 days post-vaccination onwards was 72.4% (95%CI 20.9; 94.4) and from 14 days onwards 85.2% (95%CI 41.1; 100.0).

**Table 2.**
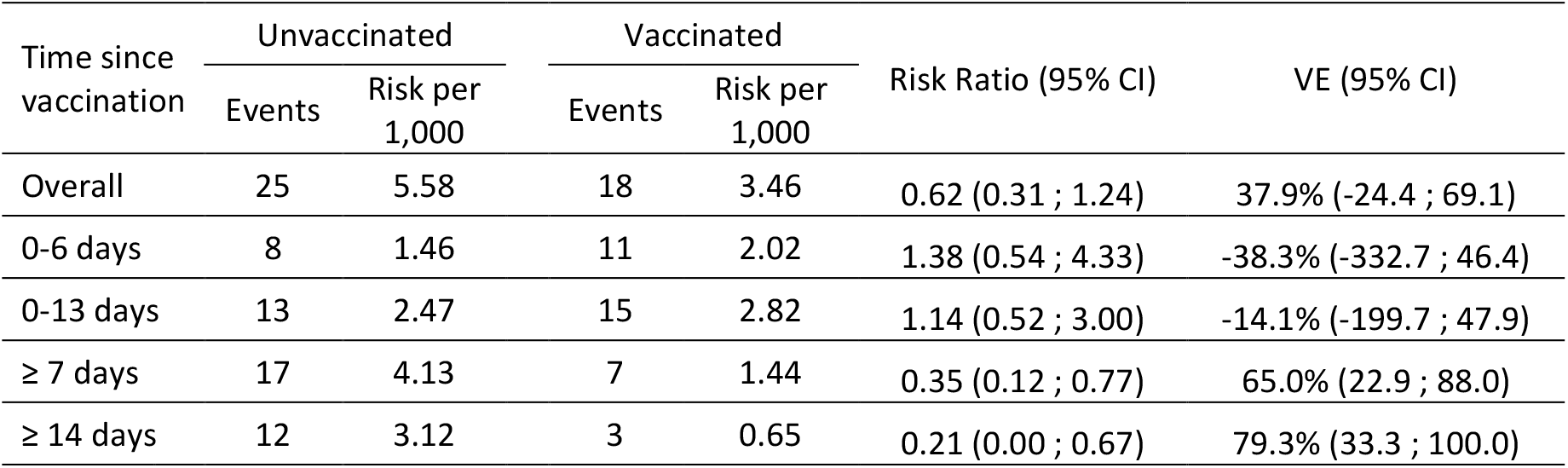
Number of events, estimated risk, risk ratios, vaccine effectiveness (VE) and 95% confidence intervals (95% CI), overall and by time since vaccination, of one dose of MVA-BN vaccine

**Table 3.** Number of events, estimated risk, risk ratios, vaccine effectiveness (VE) and 95% confidence intervals (95% CI), overall and by time since vaccination, of one dose of MVA-BN vaccine among individuals under 50 years of age

| Time since vaccination | Unvaccinated |  | Vaccinated |  | Risk Ratio (95% CI) | VE (95% CI) |
| --- | --- | --- | --- | --- | --- | --- |
|  | Events | Risk per 1,000 | Events | Risk per 1,000 |  |  |
| Overall | 22 | 5.58 | 14 | 3.00 | 0.54 (0.25 ; 1.03) | 46.3 (-3.4 ; 75.4) |
| 0-6 days | 7 | 1.44 | 9 | 1.86 | 1.29 (0.48 ; 3.93) | -29.2 (-292.8 ; 51.9) |
| 0-13 days | 11 | 2.34 | 12 | 2.52 | 1.08 (0.44 ; 2.52) | -7.8 (-151.7 ; 56.0) |
| $\geq 7$ days | 15 | 4.15 | 5 | 1.15 | 0.28 (0.06 ; 0.79) | 72.4 (20.9 ; 94.4) |
| $\geq 14$ days | 11 | 3.26 | 2 | 0.48 | 0.15 (0.00 ; 0.59) | 85.2 (41.1 ; 100.0) |

The sensitivity analysis using the full eligible population and Poisson regression obtained similar results than the main analysis (Supplement Table S2). Supplement Figure S1 depicts the person-days of follow-up and the distribution of cases and incidence rates by vaccination status throughout the study period. Overall VE during the study period was 43% (95%CI 7; 65). No vaccine effect was observed during the first 6 and 13 days post-vaccination, respectively. From 7 days onwards, VE was 68% (95%CI 35; 84), and from

14 days onwards the VE increased up to 76% (95%CI 39; 90).

## Discussion

Using a matched cohort study design of population receiving HIV-PrEP, we have estimated that the administration of one dose of MVA-BN vaccine reduces the risk of MPXV infection by 65% from 7 days post-vaccination and by 79% from 14 days post-vaccination. These results were similar in a sensitivity analysis using an alternative analytical approach, and in secondary analysis restricted to population under 50 years of age. These results confirm that MVA-BN vaccination is an effective prevention tool in a population at high-risk of MPXV infection, at least shortly after vaccine administration, with the last detected MPXV infection at around 2 months of follow-up. Since the pre-exposure vaccination campaign began when the incidence of mpox started to decrease in Spain, and has remained very low since, the effectiveness at longer times since vaccination could not be assessed.

In our study, the risk of MPXV infection in the immediate days after vaccination, particularly in the first 6 days, was higher (though not statistically significant) in the vaccinated group. This could be explained by some misclassification of post-exposure vaccines as pre-exposure (see limitations section) or because the study groups are different regarding variables we were not able to collect (e.g., sexual behavior). Since most MPXV infections will initiate symptoms within 5-7 days of exposure [27,28], it is expected that estimates of VE after 7 or 14 days of vaccination are no longer affected by misclassification of vaccine indication. On the other hand, if a higher background risk in the vaccinated truly existed, our results would indicate that MVA-BN vaccines offer protection even considering differences in MPXV infection risk.

The majority of previous studies have reached similar or higher estimates to the ones in our study. Two studies based on aggregated data conducted in the United States [16,17] in males 18-49 years old at risk of mpox, found a risk of MPXV infection more than 7 times higher in the unvaccinated compared to the vaccinated, which would be equivalent to an 86% protection conferred by the vaccine. One study in the United Kingdom [15] using the screening method [29] found a vaccine effectiveness of 78% starting 14 days after a single dose. Two studies in the United States, using case-control designs, have provided discordant estimates of VE with one dose of 75% [19] or 36% [18](72% and 41% in immunocompetent individuals, respectively), likely due to differences in the selection criteria for both cases and controls. Both studies also estimated VE of full vaccination with two doses of MVA-BN vaccine at 86% and 66%, respectively [18,19].

Sagy et al. have conducted the only cohort study currently available in the literature to estimate the effectiveness of mpox vaccination, though it included both of pre- and post-exposure vaccinations [20]. The study population were males with high-risk practices including, as in our study, men who were HIV-PrEP users but, additionally, those living with HIV and recently diagnosed with one or more STI. The study estimated a vaccine effectiveness of 86%, which however is probably over-estimated due to methodological limitations in the study, such as failure to identify equivalent time zero for the vaccinated and unvaccinated groups, leading to important confounding by calendar time [21], and the exclusion of individuals vaccinated after 26 September 2022 (presumably also of their unvaccinated follow-up time), ignoring immortal time bias in observational studies [30]. To avoid mis-specification of time zero, we emulated a target trial, as described in the literature [30,31] and as widely used in the highest quality studies on effectiveness of COVID-19 vaccines [15,32,33].

The main strengths of our study are, first, the careful specification of the start and end of the individual follow-up time and the dynamic matching of vaccinated and unvaccinated individuals to account for the time-changing baseline risk in the context of the mpox outbreak. Secondly, the availability of the indication of the vaccine allowed to specifically estimate its effectiveness administered pre-exposure, although we cannot rule out certain misclassification due to non-disclosure when attending vaccination or data collection errors. Finally, the choice of individuals on HIV-PrEP as study population has higher chances to result in a group at risk of MPXV infection with rather homogenous sexual practices and behavior, compared to studies including diverse population groups. Moreover, because MVA-MB vaccine was actively recommended in this group (in some regions even via SMS advices), we ensure that all participants had the opportunity to be vaccinated, especially since it is a group with good demonstrated access to the health-care system. This may have decreased the chance of confounding by preferential vaccine uptake in groups with different risk of exposure and/or different probability of accessing health care services when showing symptoms.

Our study has some limitations. First, even we identified over 10,000 eligible individuals, with more than 700 MPXV diagnosed infections, a limited number of events were detected in the vaccinated group, possibly due to the vaccination starting when the epidemic curve was already decreasing, on top of the protective effect of the vaccine. This has resulted in wide confidence intervals, especially in the secondary analyses. Moreover, we were not able to estimate the effectiveness of two doses of MVA-BN vaccine since all MPXV infections were registered in the first two months of follow-up, when no second doses had been administered. The lack of hospitalization or death events among the study sample prevented any estimate of effectiveness against more severe outcomes of MPXV infection.

Second, we lacked any information on risk practices and behaviours, and it is possible that individuals seeking MVA-BN vaccination were those with higher risk or, contrarily, those more preoccupied with preventive measures in general, with an unknown net effect in the VE estimate. Also, it is possible that sexual behaviours could increase following vaccination, or that people seek vaccination after a particular risk situation or a risk contact with an infected person, even if not disclosing that at the time of attending vaccination. Both situations would decrease the estimated benefit of the vaccine.

Third, we also did not have good quality information about smallpox vaccine doses administered during childhood. However, the VE for those under 50 years, for whom childhood smallpox vaccination was rare [34], yielded similar results than the main analysis, suggesting that the overall VE estimates are not significantly biased by not accounting for that variable in the analysis.

Finally, regarding the generalizability, our study was restricted to men receiving HIV-PrEP and our estimates may not be valid for the general population or for immunocompromised individuals.

## Conclusion

The administration of one dose of MVA-BN vaccine pre-exposure reduces the risk of MPXV infection in individuals on HIV-PrEP, at least shortly after vaccination. The results indicate that vaccination is an important tool for prevention and control of mpox during an outbreak. However, more studies are needed to evaluate the protection conferred by two doses of MVA-BN vaccines and the duration of the protection.

## Data Availability

The data that support the findings of this study will not be made publicly available due to data protection. Further information including the procedures for obtaining and accessing data could be obtained upon request from the corresponding author.

## Acknowledgments

The authors of the study would like to thank all the workers of the epidemiological departments, vaccination programmes, laboratory centers, and the National Health Service in every region who made all the data available and the vaccination campaign possible, alongside the work of the civil society organizations helping to reach the individuals for whom the vaccine was recommended.

## Footnotes

SM, AD, AL, VH and CO conceived the idea and elaborated the study protocol. AD, VH, CO, TV and SM coordinated the data collection. Authors from public health administrations in the Autonomous Regions were in charge of implementation of the protocol for data extraction and sharing. MF, TV and SM were in charge of data curation. MF carried out the statistical analysis and wrote the first draft of the manuscript, under the supervision of SM and with the support from AD and VH. MF is the guarantor. All authors were involved in the interpretation of study results and critically reviewed the manuscript content.

## Patient and public involvement

This study uses anonymized databases collected by the Autonomous Regions in Spain. The results of the study will be shared with the civil society organizations that helped during the vaccination campaign in Spain.

## Transparency declaration

MF, VH, AD and SM affirm that the manuscript is an honest, accurate, and transparent account of the study being reported; that no important aspects of the study have been omitted; and that any discrepancies from the study as originally planned (and, if relevant, registered) have been explained.

