## Supplementary material for "Reduction in the risk of mpox infection after MVA-BN vaccination in individuals on HIV pre-exposure prophylaxis: a Spanish cohort study"

PRE-PRINT NOT PEER-REVIEWED

### Methods and results of the sensitivity analysis using Poisson regression

To test the impact of the analytical approach, we alternatively addressed the study question using incidence rates and Poisson regression to estimate incidence rate ratios (IRR) and derive vaccine effectiveness (VE = (1-IRR)*100).

The exposure was defined as receiving at least one dose of MVA-BN vaccine, and vaccination status was assigned dynamically. All individuals started their follow-up into the study on 12 July 2022 as unvaccinated, and continued at risk in that status up to the earliest of: date of MPXV infection, date of death, date of administration of a first MVA-BN vaccine dose (regardless of indication as pre or post-exposure prophylaxis) or 12 December 2022. If a first MVA-BN vaccine dose was administered as pre-exposure prophylaxis before MPXV infection, death or 12 December 2022, the individual was classified as vaccinated and started contributing time at risk in that group up to the earliest of: date of MPXV infection, date of death, or 12 December 2022. Follow-up ended with an event when the censoring reason was MPXV infection. To assess vaccine effectiveness (VE) by number of days since dose administration, time since vaccination was further split into days 0 to 6 or ≥7 post-vaccination or, alternatively, into days 0 to 13 or ≥14 post-vaccination.

We aggregated the number of person-days of follow-up and the number of events in each vaccination status category by region (n=15), epidemiological week and age-group (18-29 / 30-39 / 40-49 / ≥50)

We used Poisson regression models to estimate the incidence rate ratio (IRR) with 95% confidence interval (95%CI), using the number of events as dependent variable, vaccination status as independent variable, and the logarithm of the number of person-days as offset. We estimated the crude IRR and the adjusted IRR by epidemiological week (using restricted cubic splines with three knots), age-group and region (as categorical variables).

**Table S1**. Baseline characteristics of the initial, the eligible and the matched sample

| | Initial sample (N=10,771) | | Eligible sample (N=10,449) | | Matched sample (N=11,320) | |
|---|---|---|---|---|---|---|
| | n | % | n | % | n | % |
| Age, years | | | | | | |
| 18-29 | 2,077 | 19.3 | 2,013 | 19.3 | 1,967 | 17.4 |
| 30-39 | 4,605 | 42.8 | 4,467 | 42.7 | 5,229 | 46.2 |
| 40-49 | 2,848 | 26.4 | 2,756 | 26.4 | 3,091 | 27.3 |
| ≥ 50 | 1,241 | 11.5 | 1,213 | 11.6 | 1,033 | 9.1 |
| Childhood smallpox vaccination | | | | | | |
| Yes | 163 | 1.5 | 135 | 1.3 | 2 | 0.0 |
| No | 33 | 0.3 | 27 | 0.3 | 92 | 0.8 |
| Unknown | 10,574 | 98.2 | 10,287 | 98.4 | 11,226 | 99.2 |
| Autonomous Region | | | | | | |
| Andalusia | 1,923 | 17.8 | 1,862 | 17.8 | 1,852 | 16.4 |
| Asturias | 53 | 0.5 | 52 | 0.5 | 74 | 0.6 |
| Balearic Islands | 768 | 7.1 | 686 | 6.6 | 722 | 6.4 |
| Canary Islands | 333 | 3.1 | 325 | 3.1 | 424 | 3.7 |
| Castile and León | 103 | 1.0 | 101 | 1.0 | 32 | 0.3 |
| Castilla-La Mancha | 76 | 0.7 | 76 | 0.7 | 118 | 1.0 |
| Catalonia | 2,925 | 27.3 | 2,813 | 26.9 | 4,242 | 37.5 |
| Valencian Community | 821 | 7.6 | 812 | 7.8 | 702 | 6.2 |
| Extremadura | 87 | 0.8 | 86 | 0.8 | 54 | 0.5 |
| Galicia | 412 | 3.8 | 407 | 3.9 | 504 | 4.4 |
| Community of Madrid | 2,493 | 23.1 | 2,473 | 23.7 | 1,660 | 14.7 |
| Region of Murcia | 194 | 1.8 | 193 | 1.8 | 258 | 2.3 |
| Navarre | 57 | 0.5 | 56 | 0.5 | 88 | 0.8 |
| Basque Country | 516 | 4.8 | 498 | 4.8 | 586 | 5.2 |
| La Rioja | 10 | 0.1 | 9 | 0.1 | 4 | 0.0 |
| MVA-BN vaccination | | | | | | |
| Yes | 5,862 | 55.4 | 5,831 | 55.8 | 5,660 | 50.0 |
| No | 4,909 | 45.6 | 4,618 | 44.2 | 5,660 | 50.0 |
| MVA-BN vaccine product | | | | | | |
| IMVANEX | 449 | 7.8 | 448 | 7.7 | 340 | 6.0 |
| JYNNEOS | 3,708 | 63.2 | 3,700 | 63.4 | 3,554 | 62.8 |
| Unkown | 1,705 | 29.0 | 1,683 | 28.9 | 1,766 | 31.2 |
| MVA-BN route of administration | | | | | | |
| Intradermal (0.1 ml) | 3,756 | 64.1 | 3,745 | 65.2 | 3502 | 61.9 |
| Subcutaneous (0.5 ml) | 1,727 | 29.5 | 1,720 | 29.5 | 1,707 | 30.2 |

PRE-PRINT NOT PEER-REVIEWED

| | | | | | | |
|---|---|---|---|---|---|---|
| Unknown | | 379 | 6.4 | 366 | 6.3 | 451 | 7.9 |
| MPXV infection | | | | | | | |
| | Yes | 738 | 6.8 | 431 | 4.1 | 43 | 0.4 |
| | No | 10,033 | 93.2 | 10,018 | 95.9 | 11,277 | 99.6 |
| Mpox symptoms* | | | | | | | |
| | Yes | 723 | 98.0 | 425 | 98.6 | 43 | 100.0 |
| | No | 15 | 2.0 | 6 | 1.4 | 0 | 0.0 |
| Hospitalization* | | | | | | | |
| | Yes | 19 | 2.6 | 11 | 2.5 | 0 | 0.0 |
| | No | 719 | 97.4 | 420 | 97.5 | 43 | 100.0 |
| Admitted to ICU* | | | | | | | |
| | Yes | 0 | 0.0 | 0 | 0.0 | 0 | 0.0 |
| | No | 738 | 100.0 | 431 | 100.0 | 43 | 100.0 |
| Death* | | | | | | | |
| | Yes | 0 | 0.0 | 0 | 0.0 | 0 | 0.0 |
| | No | 738 | 100.0 | 431 | 100.0 | 43 | 100.0 |

*Proportion is over the total number of MPXV infections

PRE-PRINT NOT PEER-REVIEWED

**Table S2.** Number of events, person-days of follow-up, estimated risk, adjusted incidence rate ratio (IRR), vaccine effectiveness (VE) and 95% confidence intervals (95%CI), of one dose of MVA-BN vaccine using adjusted Poisson regression models

| Vaccination status | | Events | Person-days | Cumulative incidence (per 10,000) | Adjusted* IRR (IC95%) | VE (95%CI) |
|---|---|---|---|---|---|---|
| Unvaccinated | | 411 | 978,730 | 4.20 | Ref. | Ref. |
| Vaccinated (time since vaccination) | Overall | 20 | 552,399 | 0.36 | 0.57 (0.35 ; 0.93) | 43% (7 ; 65) |
| | 0-6 days | 11 | 34,013 | 3.23 | 1.30 (0.71 ; 2.38) | -30% (-138 ; 29) |
| | 0-13 days | 15 | 73,464 | 2.04 | 0.94 (0.55 ; 1.59) | 6% (-59 ; 45) |
| | ≥ 7 days | 9 | 514,117 | 0.18 | 0.32 (0.16 ; 0.65) | 68% (35 ; 84) |
| | ≥ 14 days | 5 | 474,666 | 0.11 | 0.24 (0.10 ; 0.61) | 76% (39 ; 90) |

*Adjusted by calendar week, age and region within Spain.

PRE-PRINT NOT PEER-REVIEWED

**Figure S1.** Time of follow-up (a), and number of events and incidence rate (b) by vaccination status and epidemiological week in the eligible population (N=10,449)

(a)

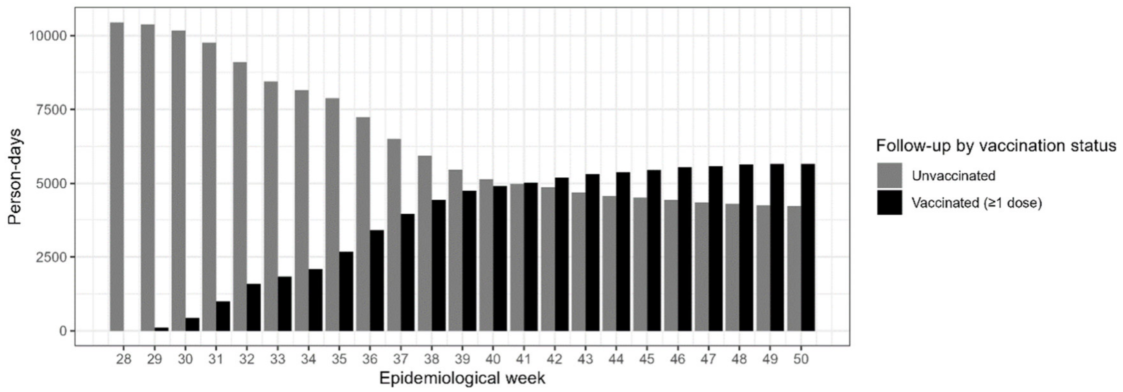


(b)

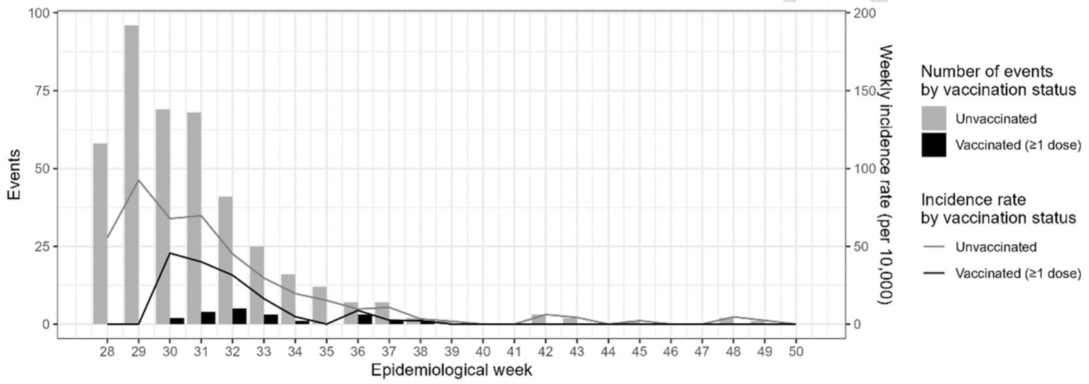
